# Time scales of human mpox transmission in the Netherlands

**DOI:** 10.1101/2022.12.03.22283056

**Authors:** Fuminari Miura, Jantien A. Backer, Gini van Rijckevorsel, Roisin Bavalia, Stijn Raven, Mariska Petrignani, Kylie E. C. Ainslie, Jacco Wallinga, the Dutch Mpox Response Team

## Abstract

Mpox has spread rapidly to many countries in non-endemic regions. After reviewing detailed exposure histories of 109 pairs of mpox cases in the Netherlands, we identified 34 pairs where transmission was likely and the infectee reported a single potential infector with a mean serial interval of 10.1 days (95% CI: 6.6–14.7 days). Pre-symptomatic transmission may have occurred in five out of eighteen pairs. These findings emphasize that precaution remains key, regardless of the presence of recognizable symptoms of mpox.

## Introduction

The current mpox outbreak was declared a public health emergency of international concern (PHEIC) by the WHO on 23 July 2022 (1). Mpox virus infection is spreading predominantly among men who have sex with men (MSM) in countries that have not reported cases of the disease previously (2).

Many key characteristics of mpox are unknown for this new mode of transmission. One such characteristic is the serial interval, defined as the time between symptom onsets of primary and secondary cases (3). Knowledge of the serial interval is key, as it informs on the reproduction number and the required intensity of control measures to stop an outbreak. Current estimates of the mean serial interval of mpox vary with a recent study estimating the mean serial interval as 5.6 days (4), while estimates have been reported of 8.5 days in the US (5), 9.5 days in UK (6), and 12.5 days in Italy (7). There is no general consensus on an estimate of mean serial interval for the current mpox outbreak, largely due to the limited availability of reliable data.

In this work, we aimed to estimate the mode, the mean and the standard deviation of serial intervals by investigating paired cases in the recent mpox outbreak in the Netherlands. We identified 109 pairs of laboratory-confirmed and notified mpox cases in the national registry with a symptom onset for the reported infector from 20 May to 3 September 2022, and a symptom onset date for the reported infectee from 24 May to 6 September 2022. All paired cases self-identified as MSM. The data were collected using contact tracing. The regional public health services that collected the data rated the reliability of self-reported symptom onset dates (into three categories: unreliable, plausible, or reliable), and assessed the likelihood of transmission between two cases (into three categories: unlikely; likely and the infectee selected an infector among several contacts; or likely and the infector is the only contact reported for the infectee). The reported symptom onset was defined for any symptom associated with mpox virus infection (8,9).

## Results

Using all 109 pairs of notified mpox cases in the national registry, the mean of observed interval between symptom onsets was 6.3 days with a standard deviation (SD) of 6.1 days (**Figure 1a**). The intervals range from -10 to 24 days, with multiple modes at 0, 4 and 8 days. The observed variation in interval duration is explained to a large extend by the likelihood of transmission between the paired cases (**Table S1** and **Methods**). After categorizing the likelihood of transmission between two cases, 34 pairs with reliable symptom onset dates were classified as likely and reported only one infector. The crude mean serial interval for those 34 pairs from all public health services was 9.4 days (SD: 6.2 days). The serial intervals range from 1 to 24 days, with a mode at 8 days. To allow for potential differences between public health services in detecting, classifying, and reporting, we used a hierarchical Bayesian framework where each public health service is treated as a random effect. The pooled mean serial interval over all public health services was 10.1 days (95% credible interval (CrI): 6.6–14.7 days) with SD of 6.1 days (95% CrI: 4.6–8.0 days) (**Figure 2**). These results were obtained using a normal distribution to describe the serial interval distribution, and similar results were obtained when repeating the analysis using a gamma distribution (mean: 10.3 days (95% CrI: 7.6–14.1 days); SD: 6.3 days (95% CrI: 4.5–9.0 days)).

**Figure 1.**
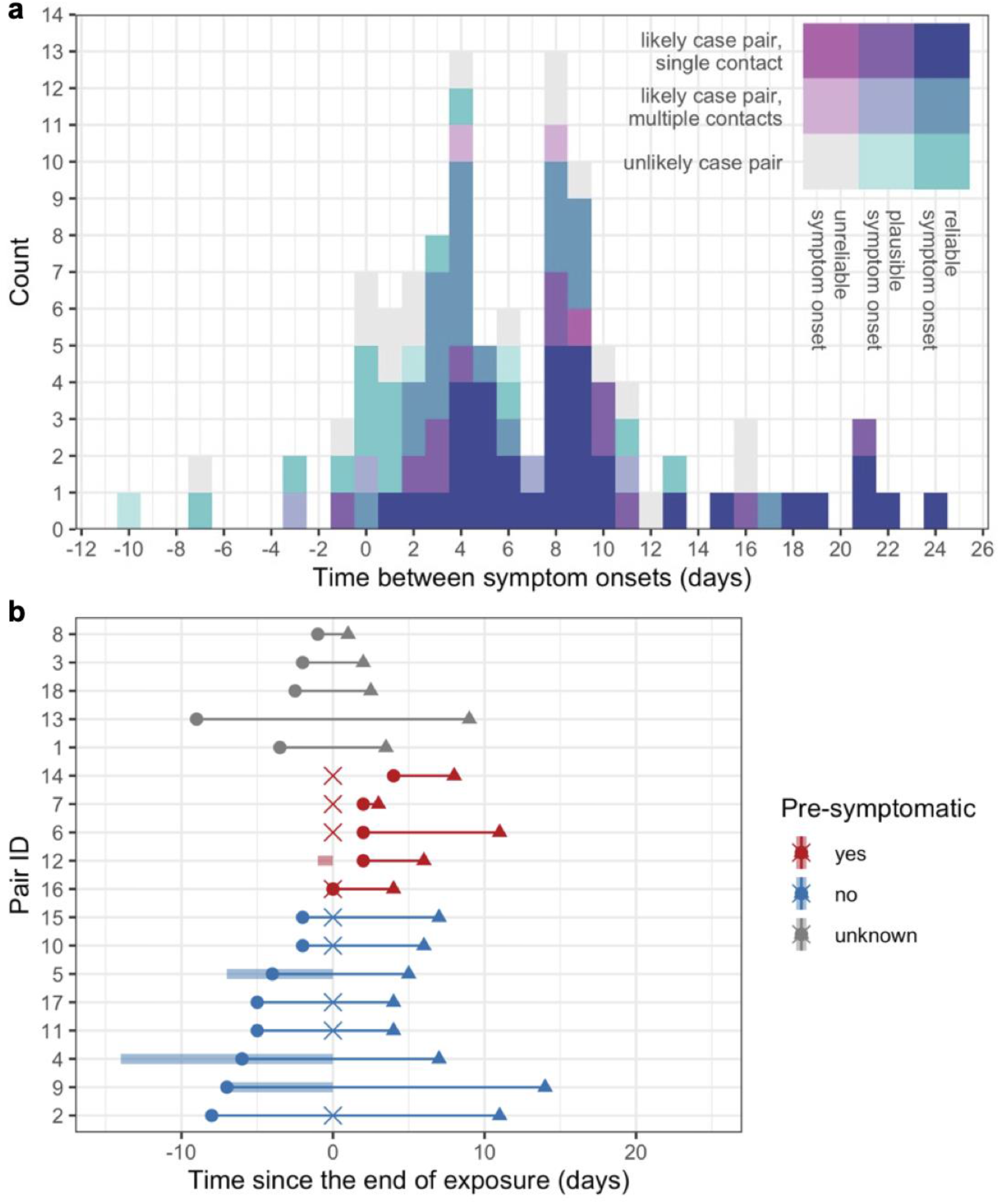
Time scale of observed transmissions. (a) Reported time differences between symptom onsets (n=109). Colors show the reliability of reporting; the reliability of self-reported symptom onset dates was rated (unreliable, plausible, and reliable) and the likelihood of transmission between two cases was categorized (contact is unlikely, contact is likely and the most plausible one among several reported contacts, contact is likely and the only contact reported for the infectee). (b) Transmission pairs notified by a single regional public health service (n=18). Circles and triangles indicate symptom onset of infectors and infectees, and the cross point is the exact date of exposure between the paired cases (if available). If the exposure date was reported as consecutive days, the time interval is visualized as a shaded bar.

**Figure 2.**
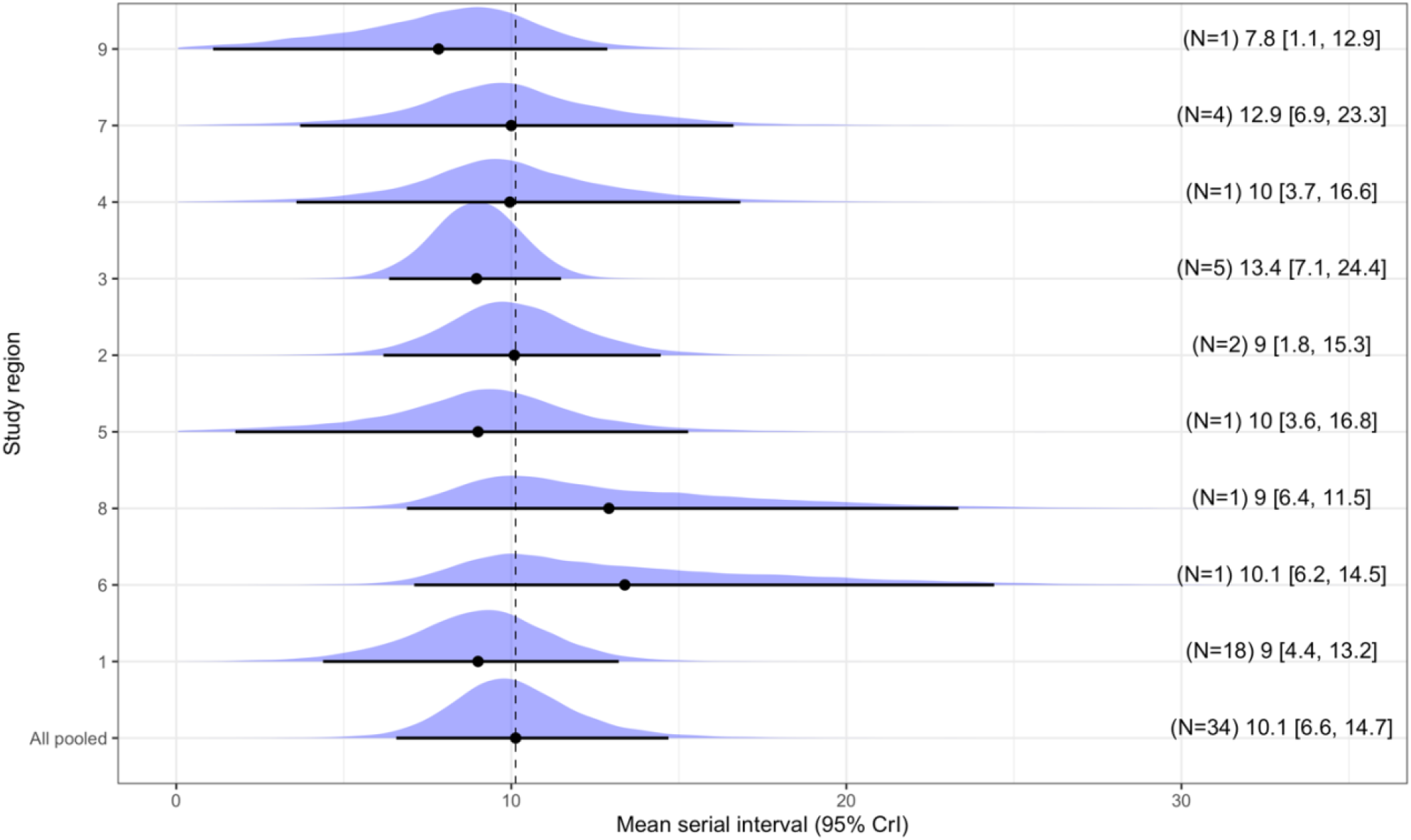
Estimated mean serial interval by regional public health service. The pooled serial interval is estimated as the average duration between symptom onset dates of a pair, incorporating random effects specific for regional public health services. Black plots represent mean values of posterior distributions, and whiskers show the 95% credible intervals.

Given the estimated pooled mean serial interval of 10.1 days (SD: 6.1 days) based on the subset of 34 pairs, we can translate the observed doubling time into the effective reproduction number *R* (i.e., the number of secondary cases produced by a typical primary case)(10). The range of values for the reproduction number *R* was estimated to be 1.3–1.6, using the average doubling time of 11.2–20.5 days during June 2022 in the Netherlands, before implementing the mass vaccination campaign (see **Supplementary materials** for a detailed derivation of the reproduction number).

For a subset of 18 pairs from a single public health service, the exposure dates were further investigated. Among the 18 pairs, 5 pairs (28%), reported contact with an infector prior to the self-reported symptom onset date of the infector, 8 pairs (44%) reported contact with an infector after the self-reported symptom onset date of the infector, and for the remaining 5 pairs (28%) the time of exposure was reported as unknown (**Figure 1b**). The close investigation of timing of exposure and symptom onset in these 18 pairs revealed that transmission can occur from 4 days before to 8 days after symptom onset of the infector, with an average duration from symptom onset to onward transmission of 2.2 days (SD: 3.9 days). Additionally, we estimated the average time between exposure and symptom onset (i.e., incubation period) for these 18 pairs (mean: 8.1 days; SD: 4.4 days), and the mean serial interval can be calculated as the sum of these mean durations, which was 10.3 days (SD: 5.9 days).

## Discussion

The present study offers empirical evidence that the average duration of the serial interval of mpox was around 10 days based on the most reliable reported transmission pairs (34 out of 109 pairs) in the Netherlands. Without strict conditions on the reliability of reporting and likelihood of transmission of infection, the mean interval between symptom onsets among all 109 pairs had a shorter duration of about 6 days.

Our observations showed that the time intervals between symptom onsets of reported pairs were highly variable and covered a wide range, without a clearly defined single mode. The wide range is consistent with variable mean values reported in earlier studies (5,7,11). These observations could be explained to a large extend by the likelihood of transmission of infection, as reported by the public health services. For the most reliable reported transmission pairs, the range of serial intervals is consistent with an infectious period that starts before and ends after the entire duration of symptoms as reported by the case. Many cases might refrain from at-risk contacts while symptomatic, either from pain or to reduce the risk of transmitting to their partners. As a consequence, transmission could occur before symptom onset and for a certain fraction of cases possibly after symptoms have disappeared. This behavioral factor gives a shorter mean and flatter distribution of the serial interval for mpox compared to smallpox, a related orthopox virus, although epidemiological characteristics for those two viruses were often considered to be comparable (12). The difference in the serial interval could be facilitated by high intensity of exposures to mpox via sexually-associated transmission routes during the current outbreak – in fact, the incubation period for human mpox and invasive smallpox infections are remarkably similar (12,13).

The frequency of transmission before a case has recognized symptoms is considerably lower than a previous report suggested, but the existence of this pre-symptomatic transmission has important implications for the outbreak control. There is a substantial risk of onward transmissions if infected individuals are unknowingly infectious. Mpox cases without any noticeable symptoms have been reported in Belgium (14), and a high viral load has been observed around the time of symptom onset among patients in the UK (15). It is likely that infected individuals are capable of sustaining a high viral load even before symptom onset thus, additional effort on monitoring and informing high-risk contacts without symptoms to adhere to temporary preventive measures may be required.

The duration of the mpox serial interval implies that the growth of the epidemic in the Netherlands was caused by the range of reproduction numbers between 1.3–1.6, which is consistent with other studies (16,17). This estimate, in turn, suggests that control measures should be sufficiently effective to prevent (1-1/1.6) × 100% = 38% of all potential secondary cases on average. Even if control measures, such as contact tracing, fail to catch the majority of contacts, they might still be highly effective in contributing to the prevention of further spread.

Our results should be interpreted with several caveats. Our analysis is restricted to cases who identified only a single infector, which may cause selection bias towards longer serial intervals because the excluded cases with multiple reported sexual contacts might have a higher frequency of sexual contact resulting in a shorter time to transmission. The analysis relies on self-reported contact history and symptom onset by notified cases. It is possible that pairs are incorrectly classified as primary-secondary infection pairs, resulting in a bias towards lower values. Heterogeneity in case finding, contact tracing, and reporting was mitigated by categorizing the pairs by the reporting public health service and treating the difference among public health services as a random effect in the analysis. Serial intervals could vary over the course of an epidemic due to right-censoring of observations induced by increasing epidemic growth, vaccination coverage, or behavioral changes due to heightened awareness. This effect is expected to be small as the study period covers both the growing and declining phases of the epidemic, and as the mass vaccination campaign started from 25 July onwards when incidence was already low (9).

In conclusion, we have estimated the mean serial interval, and showed that the current mpox outbreak in the Netherlands was driven by a moderate range of effective reproduction numbers. The estimate of the mean serial interval is conditional on the increased awareness of the disease, concomitant behaviour change, and increased immunity from natural infection and vaccination. If activity in the affected community goes back to the pre-outbreak level, and if immunity is insufficient among those at risk, there remains a risk of outbreaks or reintroduction of the virus. Our study also found that a minority of the cases might transmit infection before recognizable symptoms. This highlights that awareness remains key, regardless of the presence of recognizable symptoms, to mitigate the public health impact of resurging mpox viruses.

## Data Availability

Anonymized data and all codes used for analysis and visualization are available on Github (https://github.com/fmiura/MpxSI_2022).

## Acknowledgement

We thank the Public Health Services (GGD Amsterdam, GGD Brabant Zuid-Oost, GGD Flevoland, GGD Fryslân, GGD Gelderland-Midden, GGD Gelderland-Zuid, GGD Gooi en Vechtstreek, GGD Groningen, GGD Haaglanden, GGD Hart voor Brabant, GGD Hollands Noorden, GGD Kennemerland, GGD regio Utrecht, GGD Rotterdam-Rijnmond, GGD West-Brabant, GGD Zaanstreek-Waterland, GGD Zuid-Limburg, GGD Zuid-Holland-Zuid) for their effort to collect the epidemiological data.

## Dutch Mpox Response Team

Birgit van Benthem, Diederik Brandwagt, Hanna Bos, Colette van Bokhoven-Rombouts, Lian Bovée, Chantal P. Rovers, Brigitte van Cleef, Alje P. van Dam, Rik van Dael, Annemiek A. van der Eijk, Pauline Ellerbroek, Catharina van Ewijk, Eelco Franz, Corine GeurtsvanKessel, Joke van der Giessen, Hannelore Götz, Josette M.W. Häger, Susan van den Hof, Elske Hoornenborg, Putri Hintaran, Jorgen de Jonge, Rosa Joosten, Marion Koopmans, Kevin Kosterman, Jente Lange, Tjalling Leenstra, Daisy Ooms, Danielle Oorsprong, Eline Op de Coul, Demi Reurings, Gini van Rijckevorsel, Gregorius J. Sips, Sacha F. de Stoppelaar, Albert Vollaard, Bettie Voordouw, Harry Vennema, Henry J.C. de Vries, Karin Ellen Veldkamp, Klaartje Weijdema, Geert Westerhuis, Margreet J.M. te Wierik, Matthijs R.A. Welkers, Toos Waegemaekers, Jacco Wallinga, Paul Zantkuijl.

## Author contributions

Conceptualization: FM JW.

Data curation: FM GvR RB SR MP JW.

Formal analysis: FM JB JW.

Investigation: FM JW.

Methodology: FM JB JW.

Software: FM JB JW.

Validation: FM JB JW.

Visualization: FM JB JW.

Writing – original draft: FM JW.

Writing – review & editing: FM JB GvR RB SR MP KA JW.

## Supplementary Materials for

### Supplementary Text

#### Epidemiological data

From 21 May 2022, all suspected and confirmed cases of mpox in the Netherlands were to be notified to the regional public health services. Those cases reported their date of symptom onset, potential sources of exposure, and if known, their most likely infector. There were 109 case pairs with available symptom onsets for both cases, as of 12 September 2022. All pairs were laboratory-confirmed, according to the national diagnostic guideline (1). Other epidemiological information is publicly available on the Dutch government webpage (2).

#### Observed interval of symptom onsets

Using the case pairs, we studied the time difference between self-reported symptom onsets of the two cases. This quantity coincides with the serial interval (i.e., time between symptom onsets of primary and secondary cases) only if the infectees correctly identified and reported their infector. The set of self-reported case pairs can also contain pairs who had been infected by another common infector (co-primary cases) or pairs who had transmitted infection to the other (primary-secondary cases) with incorrect direction of transmission.

The 109 reported case pairs were collected from 19 different regional public health services in the Netherlands. These regional public health services rated the reliability of self-reported symptom onsets by three levels (i.e., unreliable, plausible, or reliable) and categorized the likelihood of transmission (i.e., unlikely; likely and the infectee selected an infector among several contacts; likely and the infector is the only contact reported for the infectee). 34 out of 109 pairs were identified as pairs with the reliable symptom onset and likely transmission with a single contact, and those pairs were collected from 9 regional public health services (**Figure 2**).

#### Bayesian random effect model

We employed a Bayesian random-effect model to obtain the pooled mean serial interval estimate, where the random effects pertain to the reporting regional public health service. The data generating process is formulated with a two-level structure, as follows.

In the first level, the *i* th observed serial interval X_*i,k*_ reported in regional public health service *k* is assumed to follow a normal distribution with mean *μ*_*k*_ and variance *σ*^2^ specific to regional public health services. This observation process is given by:

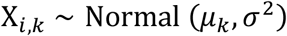

In the second level, we express the mean for each regional public health service *k* as a summation of the pooled mean 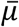 and difference *d*_*k*_. We assume that the difference *d*_*k*_ specific to each regional public health service is sampled from a normal distribution with a variance *s*^2^. This gives the following two equations:

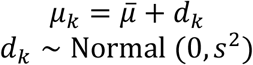

We estimated the set of parameters 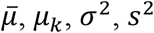 by the Markov-chain Monte Carlo (MCMC) method using Stan (3) via the {rstan} package (4). We employed weakly-informative priors Cauchy(0,10) for *σ*^2^ and *s*^2^ and improper uniform priors Uniform(−∞, ∞) for 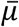 and *μ*_*k*_. The MCMC computation was performed with the default of 4 chains, 20000 samples were obtained from each chain and the first 1000 samples were discarded as warm-up iterations. The convergence of the Markov chains was assessed by r-hat diagnostic, and convergence was achieved for all parameters (3). All analyses were conducted in the R statistical programming environment version 4.0.5. (5) Reproducible codes and data are provided in the GitHub repository (https://github.com/fmiura/MpxSI_2022).

#### Growth rate and reproduction number

To illustrate the possible range of reproduction numbers in the Netherlands, we estimated doubling times using epidemic curves of confirmed cases in the Netherlands from 1 – 30 June 2022 (2). We first estimated the average exponential growth rate by performing a Poisson regression and then translated the estimated growth rate into doubling time. The estimated 95% confidence interval of doubling time in June 2022 was 11.2–20.5 days.

The estimated growth rates were then translated into reproduction numbers, using the Lotka-Euler equation (6). This requires the mean generation time τ (i.e., the mean duration between time of infection of a secondary case and of its primary case). We approximate the generation time distribution up to second order, and obtain a relationship between the reproduction number *R*, mean generation time τ, and exponential growth rate *r* is given by

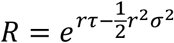

where the mean τ and variance *σ*^2^ of the generation time are identical to the pooled mean of the serial interval and the estimated variance. The range of *R* is computed based on the 95% confidence interval for the exponential growth rate *r*. The exponential growth rate *r*, in turn, is obtained from the doubling times *t*_*d*_ using the relation *r* = ln 2 / *t*_*d*_.

#### Statistical analysis of time intervals between symptom onset of self-reported case pairs

We used a mixed error-component model (7) to analyze the 109 reported time intervals. In this mixed error-component model, the reporting public health services are treated as random effects and reported reliability of symptom onsets and reported likelihood of transmission as fixed effects. The model analysis reveals statistical evidence supporting the random effects. The analysis shows there is no statistical significant effect of the reported reliability of symptom onset, but that there is a strong statistical significant effect of the reported likelihood of transmission (p<0.001) with intervals being 7.1 days (95% confidence interval: 4.1 – 10.0) shorter if transmission was reported as unlikely as compared to transmission being likely with a single contact, and a statistical significant effect (p<0.01) with intervals being 4.1 days (95% confidence interval: -0.6 – 6.0) shorter if there were multiple contacts reported as compared to transmission being likely with a single contact. The analysis reveals that, while the average interval for all reported pairs is 6.3 days, the mean interval for unlikely pairs is substantially lower, and the mean for likely transmission pairs with a single reported contact is substantially higher (**Table S1**). For the further analysis of serial intervals, we concentrate on the 34 case pairs with reliable symptom onsets and transmission between the cases was likely, with only a single reported contact, with an average of 9.4 days.

All analyses were conducted in the R statistical programming environment version 4.0.5.(5). Reproducible codes and data are provided in GitHub (https://github.com/fmiura/MpxSI_2022).

## Supplementary figures and tables

**Table S1.** Descriptive statistics of time interval between symptom onsets of self-reported case pairs, where the intervals are rated by the reporting regional health service

|  |  | Symptom onset |  |  |  |
| --- | --- | --- | --- | --- | --- |
|  |  | Reliable | Plausible | Unreliable |  |
| Transmission | Likely with a single contact | 9.4 days<br>(SD: 6.2; n=34) | 7.9 days<br>(SD: 6.3; n=12) | 9.0 days<br>(SD: NA; n=1) | 9.0 days<br>(SD: 6.1; n=47) |
|  | Likely with multiple contacts | 5.5 days<br>(SD: 3.7; n=21) | 3.8 days<br>(SD: 6.4; n=4) | 6.0 days<br>(SD: 2.8; n=2) | 5.3 days<br>(SD: 4.0; n=27) |
|  | Unlikely | 2.1 days<br>(SD: 5.2; n=14) | -0.7 days<br>(SD: 8.3; n=3) | 5.4 days<br>(SD: 6.3; n=18) | 3.6 days<br>(SD: 6.2; n=35) |
|  |  | 6.7 days<br>(SD: 6.0; n=69) | 5.7 days<br>(SD: 7.0; n=19) | 5.7 days<br>(SD: 5.9; n=21) | 6.3 days<br>(SD: 6.1; n=109) |

## References

1. WHO Director-General’s statement at the press conference following IHR Emergency Committee regarding the multi-country outbreak of monkeypox [Internet]. 2022 [cited 2022 Aug 10]. Available from: https://www.who.int/director-general/speeches/detail/who-director-general-s-statement-on-the-press-conference-following-IHR-emergency-committee-regarding-the-multi--country-outbreak-of-monkeypox--23-july-2022

2. Kraemer MUG, Tegally H, Pigott DM, Dasgupta A, Sheldon J, Wilkinson E, et al. Tracking the 2022 monkeypox outbreak with epidemiological data in real-time. Lancet Infect Dis. 2022 Jul;22(7):941–2.

3. Fine PEM. The Interval between Successive Cases of an Infectious Disease. Am J Epidemiol. 2003 Dec 1;158(11):1039–47.

4. Guo Z, Zhao S, Sun S, He D, Chong KC, Yeoh EK. Estimation of the serial interval of monkeypox during the early outbreak in 2022. J Med Virol [Internet]. 2022 Oct 21; Available from: http://dx.doi.org/10.1002/jmv.28248

5. Madewell ZJ, Charniga K, Masters NB, Asher J, Fahrenwald L, Still W, et al. Serial interval and incubation period estimates of monkeypox virus infection in 12 U.S. jurisdictions, May - August 2022 [Internet]. bioRxiv. 2022. Available from: https://www.medrxiv.org/content/10.1101/2022.10.26.22281516v1.abstract

6. Ward T, Christie R, Paton RS, Cumming F, Overton CE. Transmission dynamics of monkeypox in the United Kingdom: contact tracing study. BMJ. 2022 Nov 2;379:e073153.

7. Guzzetta G, Mammone A, Ferraro F, Caraglia A, Rapiti A, Marziano V, et al. Early Estimates of Monkeypox Incubation Period, Generation Time, and Reproduction Number, Italy, May-June 2022. Emerg Infect Dis [Internet]. 2022 Aug 22;28(10). Available from: http://dx.doi.org/10.3201/eid2810.221126

8. RIVM. RIVM LCI guideline for monkeypox [Internet]. LCI richtlijnen. [cited 2022 Jun 4]. Available from: https://lci.rivm.nl/richtlijnen/monkeypox-apenpokken

9. van Ewijk CE, Miura F, van Rijckevorsel G, de Vries HJC, Welkers MRA, van den Berg OE, et al. Monkeypox outbreak in the Netherlands in 2022: public health response, epidemiological and clinical characteristics of the first 1000 cases and protection of the first-generation smallpox vaccine [Internet]. bioRxiv. 2022. Available from: https://www.medrxiv.org/content/10.1101/2022.10.20.22281284v1.abstract

10. Wallinga J, Lipsitch M. How generation intervals shape the relationship between growth rates and reproductive numbers. Proc Biol Sci. 2007 Feb 22;274(1609):599–604.

11. Investigation into monkeypox outbreak in England: technical briefing 1 [Internet]. Gov.uk. [cited 2022 Aug 4]. Available from: https://www.gov.uk/government/publications/monkeypox-outbreak-technical-briefings/investigation-into-monkeypox-outbreak-in-england-technical-briefing-1

12. Reynolds MG, Yorita KL, Kuehnert MJ, Davidson WB, Huhn GD, Holman RC, et al. Clinical manifestations of human monkeypox influenced by route of infection. J Infect Dis. 2006 Sep 15;194(6):773–80.

13. Miura F, van Ewijk CE, Backer JA, Xiridou M, Franz E, Op de Coul E, et al. Estimated incubation period for monkeypox cases confirmed in the Netherlands, May 2022. Euro Surveill [Internet]. 2022 Jun;27(24). Available from: http://dx.doi.org/10.2807/1560-7917.ES.2022.27.24.2200448

14. De Baetselier I, Van Dijck C, Kenyon C, Coppens J, Michiels J, de Block T, et al. Retrospective detection of asymptomatic monkeypox virus infections among male sexual health clinic attendees in Belgium. Nat Med [Internet]. 2022 Aug 12; Available from: http://dx.doi.org/10.1038/s41591-022-02004-w

15. Adler H, Gould S, Hine P, Snell LB, Wong W, Houlihan CF, et al. Clinical features and management of human monkeypox: a retrospective observational study in the UK. Lancet Infect Dis [Internet]. 2022 May 24; Available from: http://dx.doi.org/10.1016/S1473-3099(22)00228-6

16. Du Z, Shao Z, Bai Y, Wang L, Herrera-Diestra JL, Fox SJ, et al. Reproduction number of monkeypox in the early stage of the 2022 multi-country outbreak. J Travel Med [Internet]. 2022 Aug 25; Available from: http://dx.doi.org/10.1093/jtm/taac099

17. CDC. Technical Report 4: Multi-National Monkeypox Outbreak, United States, 2022 [Internet]. Centers for Disease Control and Prevention. 2022 [cited 2022 Nov 2]. Available from: https://www.cdc.gov/poxvirus/monkeypox/cases-data/technical-report/report-4.html

## References

1. RIVM. RIVM LCI guideline for monkeypox [Internet]. LCI richtlijnen. [cited 2022 Jun 4]. Available from: https://lci.rivm.nl/richtlijnen/monkeypox-apenpokken

2. National Institute for Public Health and the Environment (RIVM): Monkeypox [Internet]. [cited 2022 Aug 11]. Available from: https://www.rivm.nl/en/monkeypox

3. Stan Development Team. Stan Modeling Language Users Guide and Reference Manual, ver 2.26.1 [Internet]. stan-dev.github.io. [cited 2022 Oct 20]. Available from: https://mc-stan.org

4. Stan Development Team. RStan: the R interface to Stan [Internet]. 2020. Available from: http://mc-stan.org/

5. R Core Team. R: A Language and Environment for Statistical Computing [Internet]. Vienna, Austria: R Foundation for Statistical Computing; 2020. Available from: https://www.R-project.org/

6. Wallinga J, Lipsitch M. How generation intervals shape the relationship between growth rates and reproductive numbers. Proc Biol Sci. 2007 Feb 22;274(1609):599–604.

7. Venables WN, Ripley BD. Modern Applied Statistics with S. Springer New York; 12 p.

